# Dynamics of SARS-CoV-2 Seroprevalence in a Large US population Over a Period of 12 Months

**DOI:** 10.1101/2023.10.20.23297329

**Authors:** Maria Karkanitsa, Yan Li, Shannon Valenti, Jacquelyn Spathies, Sophie Kelly, Sally Hunsberger, Laura Yee, Jennifer A. Croker, Jing Wang, Andrea Lucia Alfonso, Mondreakest Faust, Jennifer Mehalko, Matthew Drew, John-Paul Denson, Zoe Putman, Parinaz Fathi, Tran B. Ngo, Nalyn Siripong, Holly Ann Baus, Brian Petersen, Eric W. Ford, Vanathi Sundaresan, Aditya Josyula, Alison Han, Luca T. Giurgea, Luz Angela Rosas, Rachel Bean, Rani Athota, Lindsay Czajkowski, Carleen Klumpp-Thomas, Adriana Cervantes-Medina, Monica Gouzoulis, Susan Reed, Barry Graubard, Matthew D. Hall, Heather Kalish, Dominic Esposito, Robert P. Kimberly, Steven Reis, Kaitlyn Sadtler, Matthew J Memoli

**Author notes:** These authors contributed equally (Maria Karkanitsa, Yan Li, Shannon Valenti, Kaitlyn Sadtler, Matthew J Memoli).

## Abstract

Due to a combination of asymptomatic or undiagnosed infections, the proportion of the United States population infected with SARS-CoV-2 was unclear from the beginning of the pandemic. We previously established a platform to screen for SARS-CoV-2 positivity across a representative proportion of the US population, from which we reported that almost 17 million Americans were estimated to have had undocumented infections in the Spring of 2020. Since then, vaccine rollout and prevalence of different SARS-CoV-2 variants have further altered seropositivity trends within the United States population. To explore the longitudinal impacts of the pandemic and vaccine responses on seropositivity, we re-enrolled participants from our baseline study in a 6- and 12-month follow-up study to develop a longitudinal antibody profile capable of representing seropositivity within the United States during a critical period just prior to and during the initiation of vaccine rollout. Initial measurements showed that, since July 2020, seropositivity elevated within this population from 4.8% at baseline to 36.2% and 89.3% at 6 and 12 months, respectively. We also evaluated nucleocapsid seropositivity and compared to spike seropositivity to identify trends in infection versus vaccination relative to baseline. These data serve as a window into a critical timeframe within the COVID-19 pandemic response and serve as a resource that could be used in subsequent respiratory illness outbreaks.

## INTRODUCTION

More than three years have passed since the World Health Organization (WHO) identified Severe Acute Respiratory Syndrome Coronavirus-2 (SARS-CoV-2) as an emerging threat (*1*). While the emergency status for COVID-19 declared by the Centers for Disease Control and Prevention (CDC) and the WHO was lifted, the disease continues to impact on the lives of people around the world. The SARS-CoV-2 pandemic resulted in an estimated 6.46 million deaths worldwide, with an excess mortality of 14.91 million between January 2020 and December 2021 (*2*). While testing capabilities increased exponentially from the beginning months of the pandemic, the limitations of clinical polymerase chain reaction (PCR) testing made estimating percentages of those exposed or immune to SARS-CoV-2 within the general population a challenge (*3–6*). Studies to better understand exposure and immunity to SARS-CoV-2 during the pandemic period can provide information that can be used to evaluate pandemic response and provide guidance for future pandemics’ threats. Due to the wide range of COVID-19 symptoms from severe to non-existent, estimating seropositivity has allowed us to begin to identify the extent of asymptomatic infection, which significantly contributed to spread of the virus from patients who were unaware (*7–10*). In the baseline timepoint of our serosurvey, in July 2020, we showed that the rate of undiagnosed COVID seropositivity was five-fold higher than the infection diagnosis rate (*11*).

In addition to the inaccuracies that come with SARS-CoV-2 PCR testing, detection of long-term immunity to SARS-CoV-2 varies heavily on detection of antibodies that recognize the proteins making up the virus. Detection of potential humoral immunity against SARS-CoV-2 has largely relied on presence of antibodies that recognize and bind to the Spike (S) protein, including those that recognize the Spike receptor binding domain (RBD) (*12, 13*). In particular, most testing focuses on Immunoglobulin G (IgG) due to their longevity in circulation (*14*). Since our early measurements of seroprevalence in the first half of 2020, the SARS-CoV-2 pandemic evolved with the wide distribution of vaccines and the evolution of new variants that triggered new waves of infection. Through the duration of this serosurvey, four variants of concern consecutively dominated infections: B.1.1.7 (termed Alpha at the time), B.1.351 (Beta), P.1 (Gamma), and B.1.617.2 (Delta) (*15–20*). In addition to the antibody variability elicited by infection with each variant, there were further complications due to mutations of concern prevalent across multiple variants, which were projected to have severe impacts on antibody neutralization activity (*21–25*). One such mutation, the E484K mutation, was identified in the RBD and was shown to diminish antibody binding both in vaccinated and D614G convalescent samples (*26–28*). While authorized vaccines focused solely on induction of anti-S antibodies, SARS-CoV-2 infection can result in production of antibodies against other proteins that constitute the SARS-CoV-2 virus (*29, 30*). As a result, antibodies against the nucleocapsid (N) protein of the SARS-CoV-2 virus are now often used as a marker of prior infection independent of vaccination (*31, 32*). Despite this, it is well established that antibodies against nucleocapsid wane with time, with studies suggesting that anti-nucleocapsid antibody levels begin to decline within 5-15 months of infection while anti-Spike antibody levels remain high (*33, 34*). As a result, accurate identification using nucleocapsid testing would require regular antibody testing at multiple timepoints to verify prior infection without a previous positive PCR test.

While increased availability of testing has allowed for easy tracking of SARS-CoV-2 infection, the large number of undetected or unreported SARS-CoV-2 infections, the rise of different variants, and the widespread vaccine rollout has made characterizing seropositivity due to natural infection, vaccination, or both extremely difficult. This study aims to provide longitudinal data that could further clarify the dynamics of SARS-CoV-2 seroprevalence in the United States over the course of the pandemic, accounting for prior infections, SARS-CoV-2 variants, and antibody classes in a multi-analyte longitudinal serologic study. Furthermore, this study highlights the seropositivity of US adults not only at the beginning of the pandemic, but also just before and immediately after vaccine rollout to the general population within the United States.

## RESULTS

### Attrition rates for re-enrolled participants varied within different demographic groups

Prior to collecting the two follow-up timepoints, we re-enrolled individuals from a convenience sample that was selected via quota-based sampling as described in our original publication (**Figure 1**) (*11*). 6- and 12-month enrollment occurred from September 2^nd^, 2020 – March 2^nd^, 2021 and from March 2^nd^, 2021 – December 5^th^, 2021, respectively. Of those who agreed to participate in the follow ups, 4562 (56.6%) participants from our initial serosurvey completed the 6-month follow up, and 4226 (52.44%) participants completed the 12-month timepoint (**Table 1**). A total of 2,704 participants returned samples and survey information for all three timepoints. The greatest attrition at the 12-month timepoint came from the rural group, where 79.3% fewer participants re-enrolled as compared to the urban group, where attrition was 42.35%. The lowest attrition rates at the 12-month timepoint came from the elderly (70+) population at 36.21% fewer participants, compared to the age group of 18-44 years old with 56.23% attrition.

**Figure 1.**
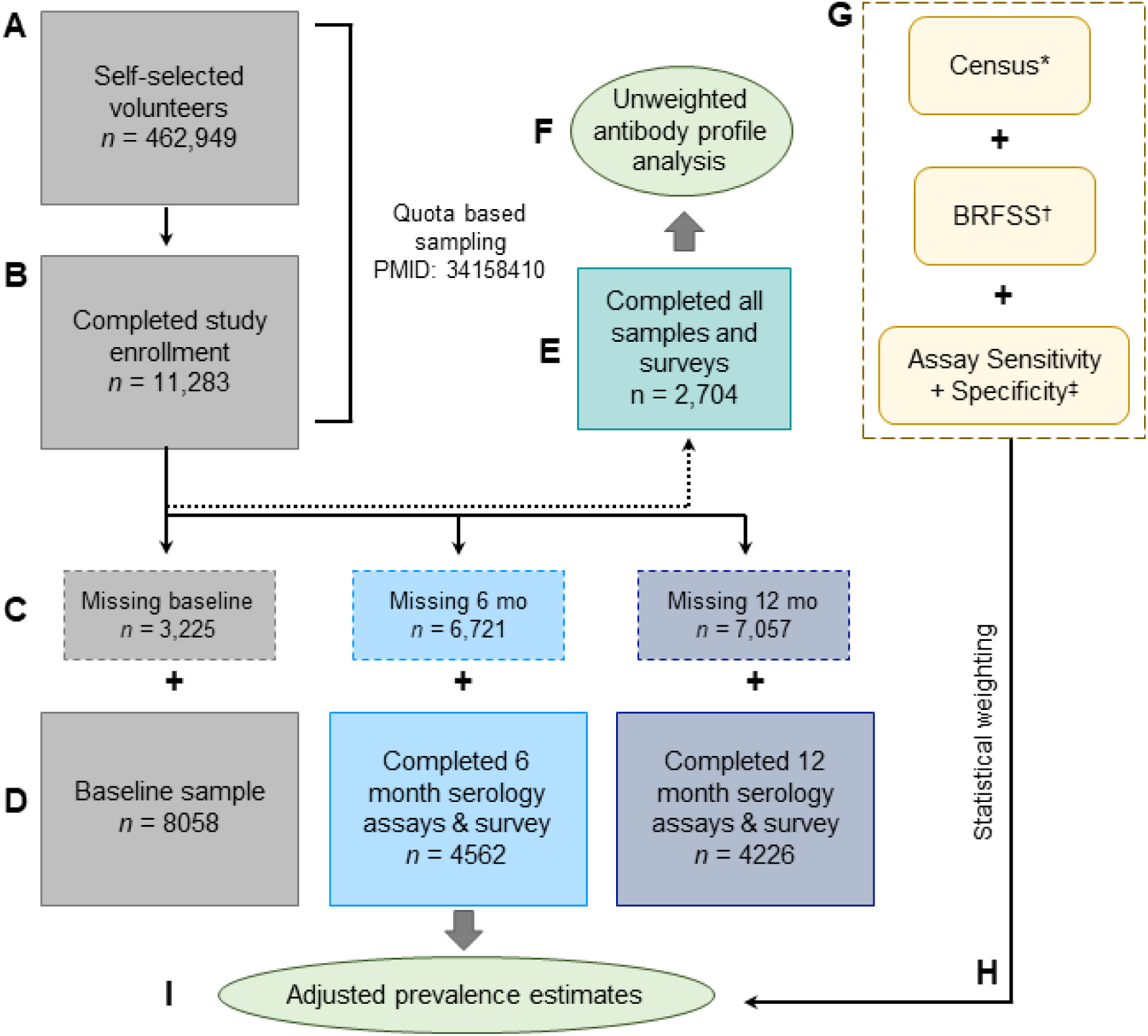
Study design and statistical workflow. (A) Initial volunteer pool from self-selected volunteers. (B) Resulting 11,283 participants from quota-based sampling generated through Census and BRFSS as previously described (*11*). (C) Participants with missing samples or survey information removed from study. (D) Participants that returned full survey and samples for each timepoint. (E) Participants that returned samples and survey information for all timepoints tested (F) Exploratory endpoint of antibody profile in unweighted analysis (within study population). (G) Factors that were fed into weighting. (H) Statistical weighting applied to different seroprevalence estimates (I) Primary endpoint of seroprevalence within timepoints. * Census = US Census Data for 2018 . ^†^ BRFSS = Behavioral Risk Factor Surveillance System (CDC) (*46*). ^‡^ Assay Sensitivity and Specificity as determined by prior studies (spike/RBD, (*6, 11*)) and evaluations of new antigens (Nucleocapsid, see Methods and Supplemental Figure 1).

**Table 1.** Participant demographics. Number (n) of people that returned full survey and blood sample within each time point. % = fraction of persons compared to the total study population. * = census or weighted BRFSS data (CDC Behavioral Risk Factor Surveillance System). Time point 0 = previously published initial survey PMID: 34158410, (*11*).

| Timepoint | n |  |  | % |  |  | BRFSS/<br>Census* | Attrition %<br>t = 0 to 12 |
| --- | --- | --- | --- | --- | --- | --- | --- | --- |
|  | 0 | 6 | 12 | 0 | 6 | 12 |  |  |
| **Total** | 8058 | 4562 | 4226 | 100 | 100 | 100 |  | 47.56 |
| **Northeast** | 1345 | 767 | 712 | 16.69 | 16.81 | 16.85 | 17.6 | 47.06 |
| **Midwest** | 1276 | 750 | 716 | 15.84 | 16.44 | 16.94 | 16.97 | 43.89 |
| **Mid-Atlantic** | 1674 | 895 | 814 | 20.77 | 19.62 | 19.26 | 16.91 | 51.37 |
| **South/Central** | 1143 | 672 | 628 | 14.18 | 14.73 | 14.86 | 15.35 | 45.06 |
| **Mountain/Southwest** | 1252 | 694 | 650 | 15.54 | 15.21 | 15.38 | 15.89 | 48.08 |
| **West/Pacific** | 1368 | 784 | 706 | 16.98 | 17.19 | 16.71 | 17.27 | 48.39 |
| **18 - 44** | 3349 | 1702 | 1466 | 41.56 | 37.31 | 34.69 | 46 | 56.23 |
| **45 - 69** | 3436 | 2072 | 1948 | 42.64 | 45.42 | 46.10 | 39.84 | 43.31 |
| **70+** | 1273 | 788 | 812 | 15.80 | 17.27 | 19.21 | 14.17 | 36.21 |
| **Male** | 4241 | 2117 | 1957 | 52.63 | 46.41 | 46.31 | 48.66 | 53.86 |
| **Female** | 3817 | 2445 | 2269 | 47.37 | 53.59 | 53.69 | 51.34 | 40.56 |
| **White** | 6243 | 3621 | 3392 | 77.48 | 79.37 | 80.27 | 73.41 | 45.67 |
| **Black** | 761 | 409 | 352 | 9.44 | 8.97 | 8.33 | 12.9 | 53.75 |
| **Other Race** | 1054 | 532 | 482 | 13.08 | 11.66 | 11.41 | 13.69 | 54.27 |
| **Hispanic/Latino** | 1281 | 682 | 593 | 15.90 | 14.95 | 14.03 | 17.06 | 53.71 |
| **Not H/L** | 6777 | 3880 | 3633 | 84.10 | 85.05 | 85.97 | 82.94 | 46.39 |
| **Urban** | 6923 | 4316 | 3991 | 85.91 | 94.61 | 94.44 | 93.48 | 42.35 |
| **Mostly Rural** | 1135 | 246 | 235 | 14.09 | 5.39 | 5.56 | 6.52 | 79.30 |
| **Children <18yrs in house** | 2616 | 1379 | 1185 | 32.46 | 30.23 | 28.04 | 35.81 | 54.70 |
| **No children** | 5442 | 3183 | 3041 | 67.54 | 69.77 | 71.96 | 64.19 | 44.12 |
| **High School** | 210 | 105 | 103 | 2.61 | 2.30 | 2.44 | 41.07 | 50.95 |
| **Some college** | 1113 | 605 | 528 | 13.81 | 13.26 | 12.49 | 30.88 | 52.56 |
| **College graduate** | 6735 | 3852 | 3595 | 83.58 | 84.44 | 85.07 | 28.05 | 46.62 |
| **Own** | 6058 | 3561 | 3337 | 75.18 | 78.06 | 78.96 | 66.49 | 44.92 |
| **Rent** | 1625 | 832 | 727 | 20.17 | 18.24 | 17.20 | 27.32 | 55.26 |
| **Other** | 375 | 169 | 162 | 4.65 | 3.70 | 3.83 | 6.19 | 56.80 |
| **Employed** | 5738 | 3127 | 2827 | 71.21 | 68.54 | 66.90 | 57.74 | 50.73 |
| **Other** | 1920 | 1191 | 1182 | 23.83 | 26.11 | 27.97 | 31.38 | 38.44 |
| **Unemployed** | 400 | 244 | 217 | 4.96 | 5.35 | 5.13 | 10.88 | 45.75 |
| **Health Care Coverage** | 7851 | 4469 | 4142 | 97.43 | 97.96 | 98.01 | 87.85 | 47.24 |
| **No Health Care** | 207 | 93 | 84 | 2.57 | 2.04 | 1.99 | 12.15 | 59.42 |

We evaluated seroprevalence in these groups as an overall seropositivity focused on antibody reactivity to SARS-CoV-2 spike (S) protein and its receptor binding domain (RBD), which can be induced by infection, vaccination, or a combination of both. To account for potential variance in background reactivity among patients with different exposure histories, we determined seropositivity based on prevalence of both anti-S and anti-RBD antibodies as previously described (*11*).

### Seropositivity in participants of all three timepoints shows positive correlation between anti-Spike and anti-RBD antibodies across all antibody classes

In evaluating the samples received by participants in all three timepoints outlined in the study (**Fig 2A**), we identified a visual correlation between Spike and RBD seropositivity across all samples probed (**Fig 2B-2G).** There was also a numerical increase in median optical density (OD) values for anti-Spike and anti-RBD antibodies, regardless of class. While the median OD values for IgG, IgM, and IgA antibodies fall below the level of detection at the 6-month timepoint, all the median thresholds for these antibodies were above the threshold at 12 months. This trend is particularly strong in IgG and IgA, but is not as pronounced in IgM anti-Spike and anti-RBD antibodies (**Fig 2C and 2F**). While IgG antibody values were less dispersed at 12 months relative to 6 months (**Fig 2B and 2E**), IgM and IgA antibodies were far more dispersed at the 12-month timepoint for both anti-Spike and anti-RBD antibodies (**Fig 2C-2D**, **Fig 2F-2G**). The median OD value for anti-nucleocapsid antibodies at 12 months was far lower than the median value for anti-RBD antibodies (**Fig 2H**).

**Figure 2.**
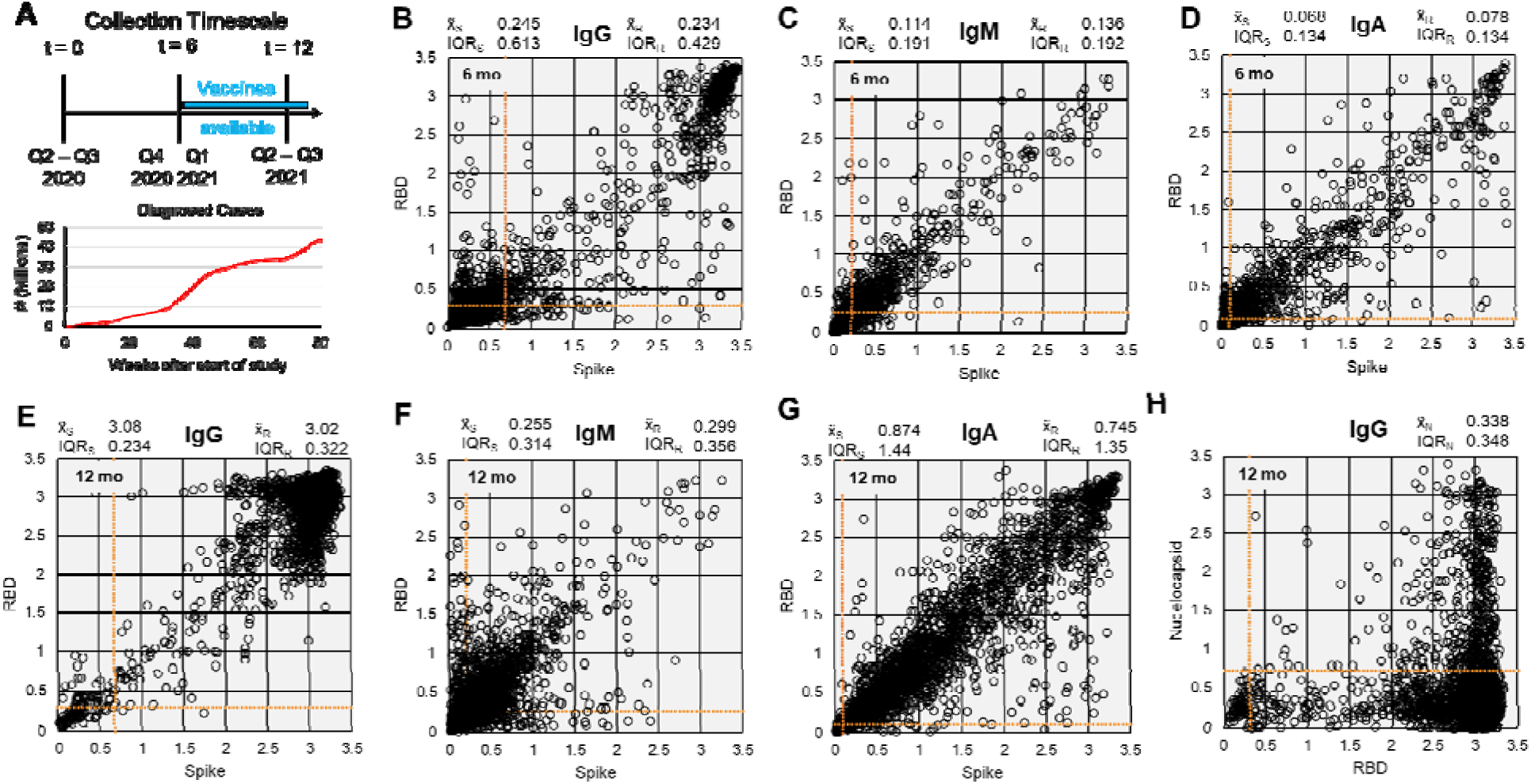
Raw serology data for participants who submitted samples for all three time points. (**A**) Sample collection timescale and cumulative count of diagnosed cases during the study period (data from CDC). Data distribution for 6-month timepoint of (**B**) IgG, (**C**) IgM, and (**D**) IgA antibodies against Spike (trimer) and RBD. (**E-H**) Data distribution for 12-month timepoint of (**E**) IgG, (**F**) IgM, and (**G**) IgA antibodies against Spike (trimer) and RBD, and (**H**) Nucleocapsid (C-terminal domain). Data displayed are raw optical density (OD) readings after background correction. Data are from 2,704 participants with samples from all 3 timepoints. x̃_S_ = median OD for Spike; x̃_R_ = median OD for RBD; x̃_N_ = median OD for Nucleocapsid; IQR = interquartile range (unweighted). Linear range of plate reader = OD 1 - 3. Orange = positivity threshold values per analyte.

Interestingly, there were a few samples at 6 months that were either Spike or RBD IgG positive, but not both (**Fig 2B**). The number of samples where this occurred, however, was visibly lower by 12 months (**Fig 2E**). The opposite occurred in IgM samples, where there are visibly more samples that are either anti-Spike or anti-RBD IgM positive (**Fig 2C**, **Fig 2F**). The same phenomenon was observed in IgA, where there were visibly more Spike-high, RBD-low samples relative to 6 months (**Fig 2D**, **Fig 2G**). At 12 months, correlation between nucleocapsid and anti-RBD-IgG antibody levels was weak, indicating that high anti-RBD IgG antibodies were likely due to vaccination and not recent infection (**Fig 2H**). There were a few samples with high anti-nucleocapsid and low anti-RBD samples. However, there were more samples that had high levels of nucleocapsid and RBD antibodies, suggesting prior infection or infection and vaccination as the source of seropositivity.

### Overall seropositivity increased with time and varied with demographic and socioeconomic factors

As expected, seropositivity was greater at 6 (prevalence: 36.2%, 95% CI: 30.2 – 43.1) and 12 months (89.3%, 95% CI: 83.8 – 96.8), respectively, when compared to our initial published estimate at baseline (4.6%, 95% CI: 2.6 – 6.5) (**Fig. 3**). At 6 months there were larger regional variations compared to 12 months, with Mid-Atlantic (44.9%, 27.2 – 64.4) and South/Central (51.5%, 33.2 – 70.6) regions displaying higher point estimates. 12-month estimates remained consistently high across all regions (82.5% - 93.5%), with the highest point estimate being in the Mid-West (93.5%, 95% CI: 87.6 – 100). As with our initial study, the youngest age group maintained the highest seroprevalence at 6 months at 43% (32.1 – 55.2) compared to the 45 – 70 (27.9%, 23.5 – 33.0) and 70+ groups (37.1%, 27.6 – 48.0). After introduction of vaccines, there was a sharp increase in seroprevalence of the 70+ age group that correlated with external vaccine uptake data, showing 98.7% seropositivity (97.3 – 100) in comparison to the 18 – 44 age group which increased less drastically, at 84.9% seropositive (73.3 – 95.5). Though males had slightly higher prevalence in our initial study, at 6 months this was shifted to women having higher prevalence than men (women 41.9%, 33.3 – 51.7; men 29.7%, 22.6 – 37.9). This pattern again inverted, with both groups increasing by 12 months (women 86.9%, 77.6 – 96.2; men 92.0%, 86.7 – 99.5). Initially, compared to White participants, Black participants had higher seroprevalence and this pattern was maintained through the 6-month timepoint with a large confidence interval (White 35.3%, 28.9 – 42.7; Black 46.0%, 23.1 – 71.2). By 12 months this pattern inverted, though again there was a large confidence interval for point estimates. Hispanic and Latino participants had similar seroprevalence compared to non-Hispanic/Latinos at 6 months, but higher seroprevalence by 12 months (Hispanic 96.7%, 93.0 – 100; Non-Hispanic 87.9%, 81.4 – 95.9). Persons that lived in rural areas versus those that lived in urban areas had similar seroprevalence at 6 months, with urban dwellers having higher overall seroprevalence at 12 months (urban 90.9%, 85.5 – 98.4; rural 64.3%, 38.9 – 86.5).

**Figure 3.**
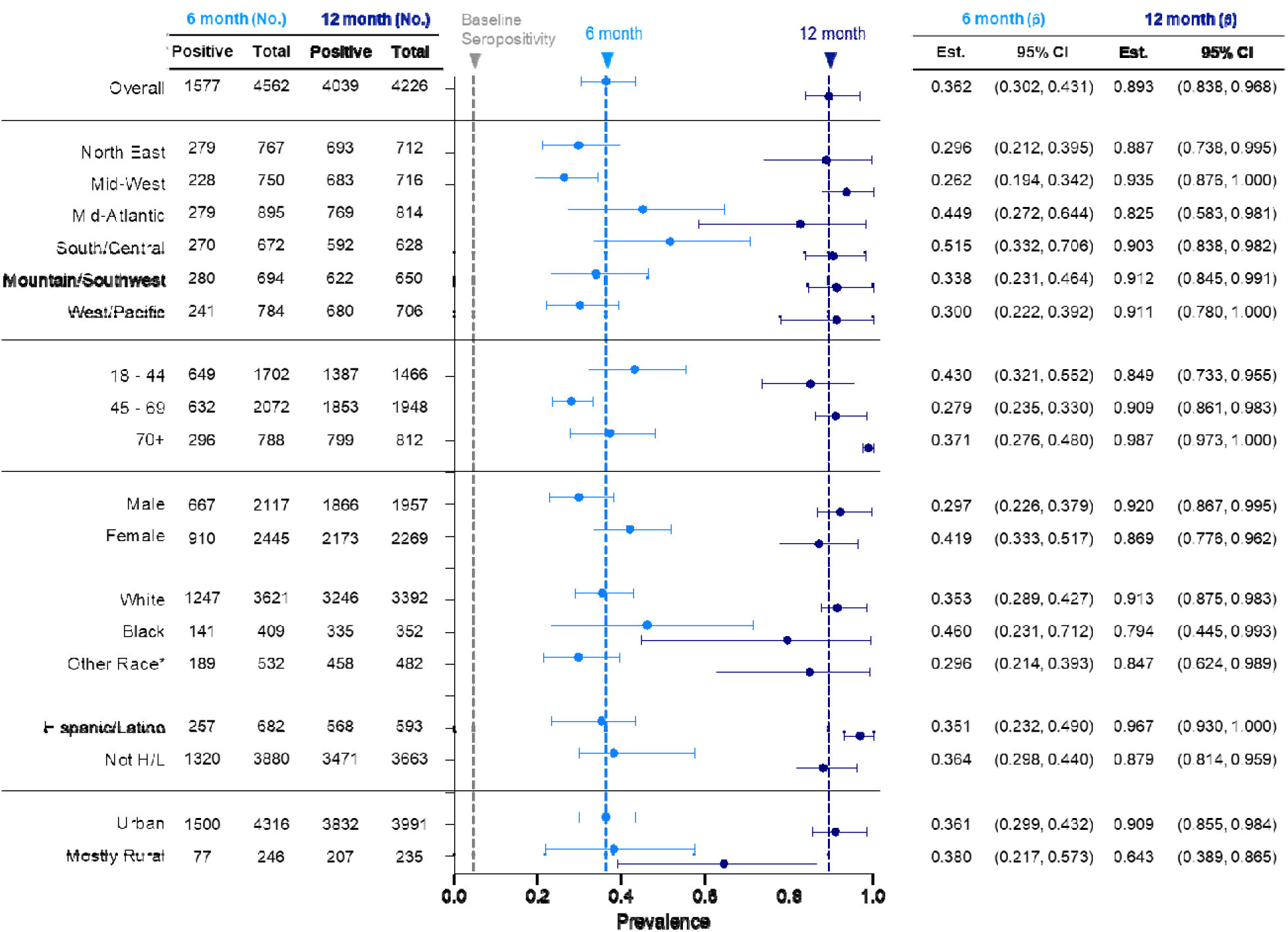
Demographics of seropositivity at 6 and 12 months after initial testing. No. = number, p̂ = proportion, CI = confidence interval. Grey vertical dashed line = initial overall seroprevalence estimate at Q3 2020. Light blue vertical dashed line = 6-month overall seroprevalence estimate. Dark blue vertical dashed line = 12-month overall seroprevalence estimate. Data are weighted point estimates for prevalence and 95% confidence intervals. Initial testing (t = 0) data available via PMID: 34158410.

In terms of socioeconomic factors, people in households with children under 18 years old had a slightly higher seroprevalence at 6 months (with children: 43.4%, 32.1 – 56.0, no children: 31.9%, 25.8 – 39.0) but this difference disappeared by 12 months after the initial study (**Fig. 4**). In our first study, seroprevalence was similar regardless of education level. By 6 months, however, seroprevalence increased in those with a high school education or less (43.8%, 27.0 – 62.6) compared to those with college or technical school experience (34.1%, 31.5 – 37.6). By 12 months, prevalence shifted with college graduates having the highest seroprevalence (95.3%, 94.2 – 100) and those with a high school education or less having the lower seroprevalence (83.3%, 65.1 – 96.8). The latter group, however, had a large range of uncertainty due to the lower number of respondents, as reflected in the confidence intervals. No difference between those who owned versus rented housing was noticed until the 12-month timepoint, when it was noted that those who owned homes had higher seroprevalence than those who rented or responded “other” (own 93.0%, 90.2 – 99.8; rent 79.5%, 65.3 – 92.0; other 80.3%, 30.8 – 100). Those that were unemployed had the highest seroprevalence point estimate at 6 months (57.9%, 31.1 – 83.1) and the lowest at 12 months (68.0%, 34.6 – 93.4) when compared to other groups.

**Figure 4.**
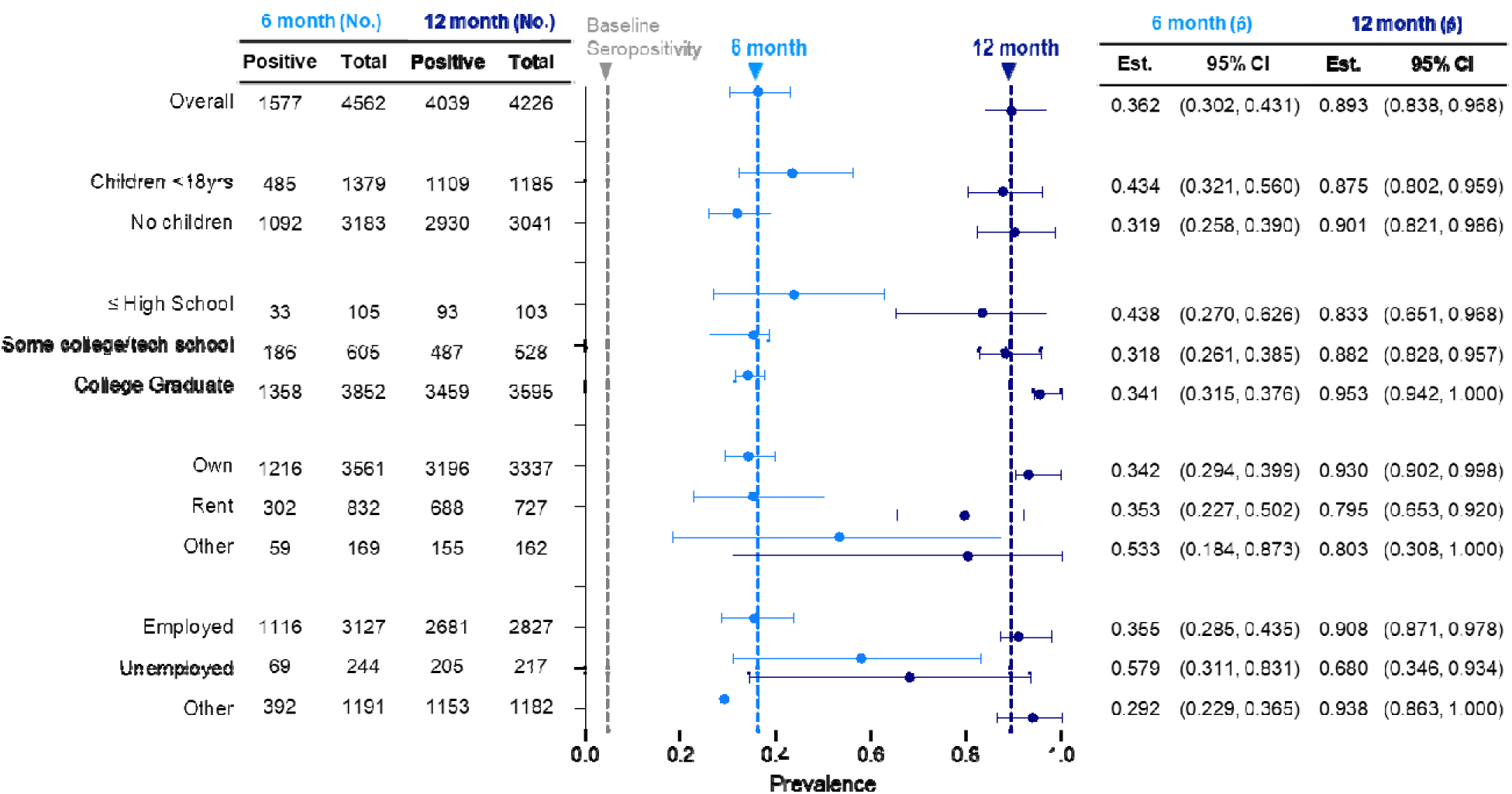
Socioeconomic factors associated with seroprevalence at 6 and 12 months after initial testing. No. = number, p̂ = proportion, CI = confidence interval. Grey vertical dashed line = initial overall seroprevalence estimate at Q3 2020. Light blue vertical dashed line = 6-month overall seroprevalence estimate. Dark blue vertical dashed line = 12-month overall seroprevalence estimate. Data are weighted point estimates for prevalence and 95% confidence intervals.

In evaluating health-related factors, we noted that the estimated seroprevalence for those without healthcare coverage was higher at 6 months relative to those with coverage, a difference which was not noticed in our initial timepoint (**Fig. 5**). In accordance with our first timepoint, those with diabetes had lower seropositivity compared to those without diabetes (23.0%, 13.9 – 34.7; 37.5%, 31.1 – 44.9). This gap decreased by the 12-month timepoint. While there was a trending decrease in those who received a flu or pneumonia shot at 6 months, there was a higher seroprevalence in those with a flu/pneumonia shot at 12 months (with shot: 95.2%, 91.6 – 100; no shot: 82.9%, 72.5 – 93.1).

**Figure 5.**
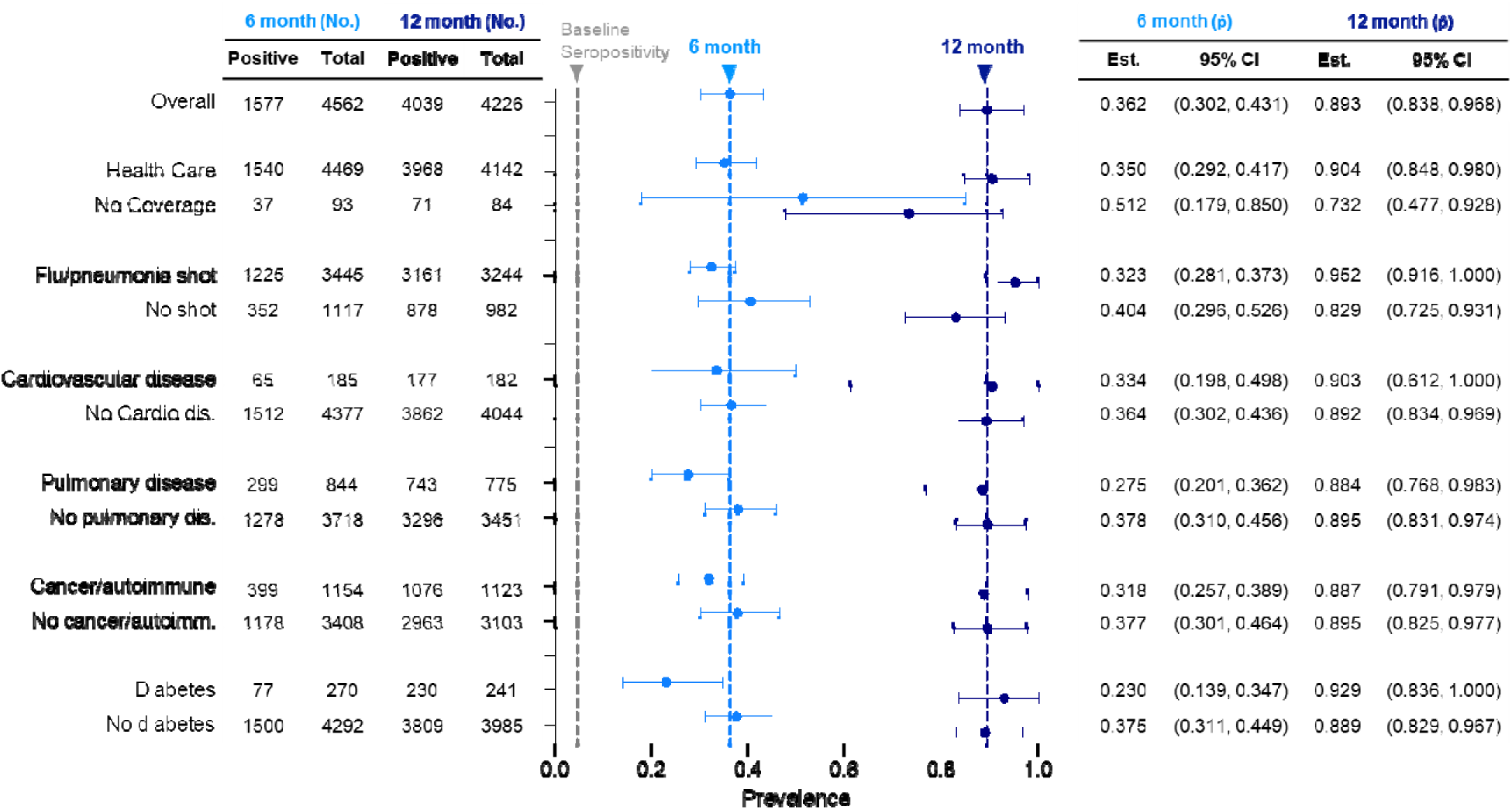
Health factors affecting seroprevalence at 6 and 12 months after initial testing. No. = number, p̂ = proportion, CI = confidence interval. Grey vertical dashed line = initial overall seroprevalence estimate at Q3 2020. Light blue vertical dashed line = 6-month overall seroprevalence estimate. Dark blue vertical dashed line = 12-month overall seroprevalence estimate. Data are weighted point estimates for prevalence and 95% confidence intervals.

### Infection-induced nucleocapsid seroprevalence varied across different demographic and socioeconomic groups

In our initial timepoint study, 4.6% of the samples were seropositive for anti-S and anti-RBD antibodies, all due to infection as vaccines had not yet been introduced. We further identified that 71.5% (unweighted) of baseline seropositive samples were nucleocapsid (Nuc) positive, with 41.5% of Nuc IgG-samples being S-IgG negative and those that were nucleocapsid negative also having lower spike IgG levels (Nuc IgG+ anti-S IgG OD = 3.52, Nuc IgG-anti-S IgG OD = 1.065; Supplemental Figure 1,2). At the 12-month timepoint, we identified that 17.3% (12.9 - 22.0) of our total samples were seropositive for anti-Nuc IgG, suggesting exposure to SARS-CoV-2 (with or without vaccination, Figure 6). Geographically, all areas had similar anti-Nuc infection-induced seroprevalence with the West/Pacific and Northeast having the lowest anti-Nuc seroprevalence (8.9%, 3.1 – 16.7; 12.7%, 6.4 – 20.7). All ages and both male and female participants had similar prevalence estimates. Black participants had the highest anti-Nuc seroprevalence estimate, albeit with a large error range (26.1%, 10.5 – 47.4). Those without children had slightly lower anti-Nuc seroprevalence compared to those with children in the household under 18 years old (14.8%, 10.3 – 19.5; 22.6, 14.3 – 32.7). When evaluating education level, college educated participants had the lowest anti-Nuc seroprevalence compared to other groups (13.0%, 10.1 – 15.6). When evaluating the effect of healthcare coverage, we found that those without healthcare had higher anti-Nuc seroprevalence compared to those with healthcare coverage (42.9%, 20.8 – 68.1; 15.6% 11.1 – 20.1). As with our 6-month timepoint, there is a lag in seropositivity for both nucleocapsid and spike within rural populations (nucleocapsid: 11.1%, 0.8 - 31.6; spike: 64.3%, 0.389 – 0.865) compared to urban populations (nucleocapsid: 17.1%, 13.1-22.6; spike: 90.9%, 0.855 – 0.984).

**Figure 6.**
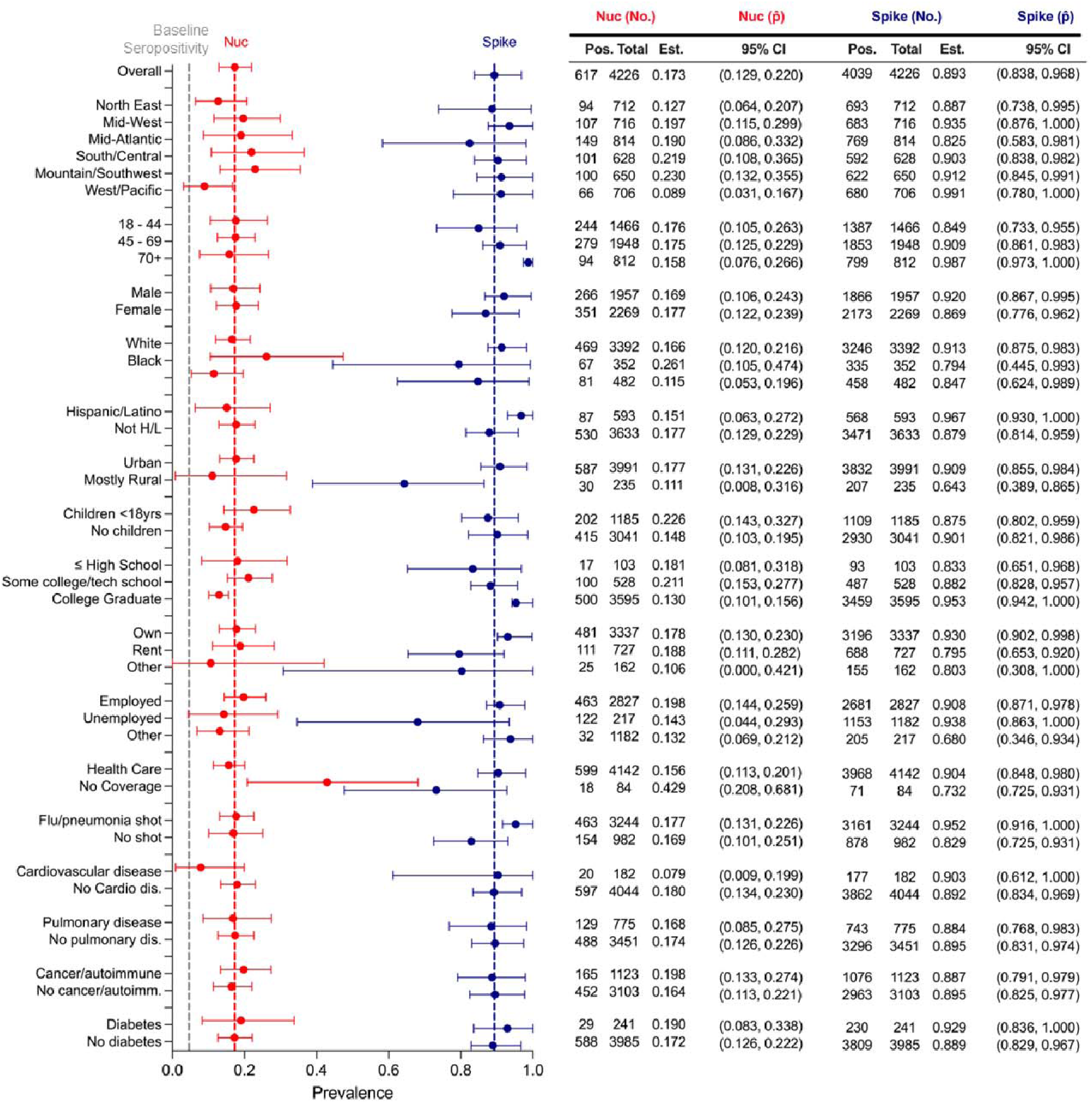
Infection-induced nucleocapsid seroprevalence at commencement of vaccine rollout. Seropositivity evaluation at 12 months after baseline testing (t = 0) after initial vaccine rollout in the United States. Red = Nucleocapsid IgG seropositivity. Dark blue = Spike/RBD IgG/IgM combined seroprevalence. *self-reported races including Asian American/Pacific Islander, Native American, and others had large confidence intervals and were combined into a single point estimate.

### Tracking antibody profile and antigen reactivity in individuals from pandemic onset to vaccine rollout

Within a subset of seropositive participants, we were able to evaluate their antibody profile over the course of the rollout of vaccination in the United States (**Fig. 7**). As previously noted, IgG antibody persistence was strong throughout all three timepoints (**Fig. 7A**). Due to introduction and availability of SARS-CoV-2 vaccines between timepoint 6 and 12 months, we observed increased seroprevalence and strong anti-RBD IgG seropositivity at 12 months among individuals from whom we had samples across multiple timepoints. IgM antibody prevalence faded between timepoints (**Fig. 7B**), with waning observed in samples between baseline (t = 0), 6-month, and 12-month timepoints. Serum IgA mirrored IgM, with fading after 0- and 6-month timepoints. However, we noted a strong induction of serum IgA antibodies at 12 months, possibly due to vaccination (**Fig. 7C**). At the 12-month timepoint, we evaluated the S-RBD seropositive individuals for nucleocapsid antibody prevalence as well as reactivity against immune-evasive mutations within the RBD, specifically an E484K mutation that appeared within the delta variant (**Fig. 7D**). In seropositive individuals, nucleocapsid prevalence was strongest in the 0-month timepoint followed by the 6- and 12-month timepoints. This is likely due both to vaccination and nucleocapsid antibody waning from prior infections (**Fig 7E**). When comparing vaccinated versus unvaccinated individuals at the 12-month timepoint, there was a significantly higher anti-Nuc IgG normalized OD in unvaccinated individuals when compared to vaccinated individuals (**Fig. 7F**). Within the vaccinated subset 12.78% of individuals had reported a prior infection during the course of the study (9.64%) or were detected as seropositive in the initial timepoint (3.14%). To further determine trends in seropositivity, we assessed correlations across all the analytes we probed for in our samples. Overall, there were similar correlations of antibody reactivity across all analytes in the different timepoints. We did note that, while nucleocapsid IgG was positively correlated with trends in Spike and RBD IgG levels in the initial timepoint, this correlation shifted negatively in timepoints 6 and 12 (**Fig. 7G – I**).

**Figure 7.**
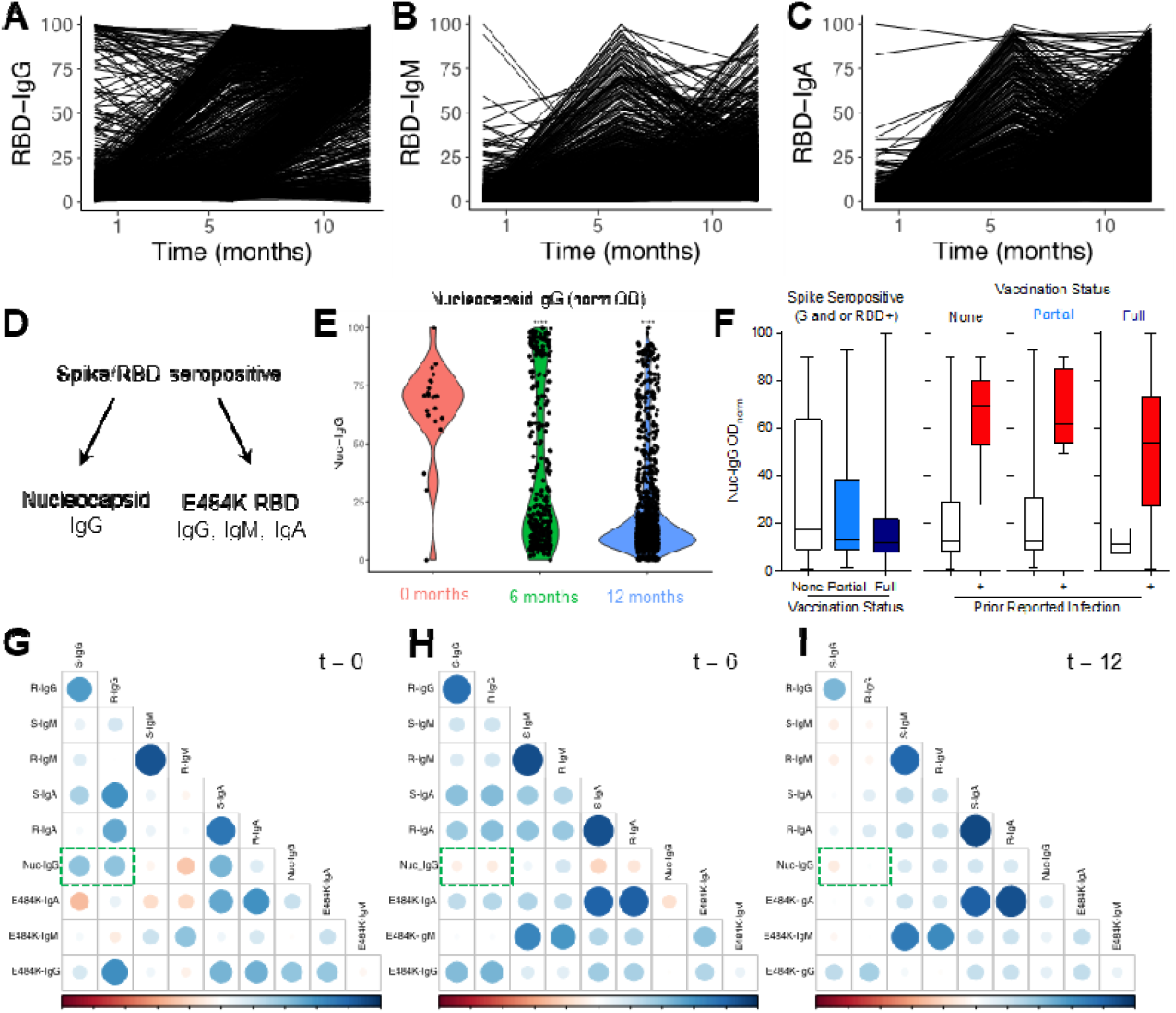
Antibody dynamics over a 12-month period at the beginning of the COVID-19 pandemic. (A) RBD IgG (B) IgM, and (C) IgA normalized (0–100) optical density (OD) in participants that returned all three samples (n = 2,704). (D) Evaluation of expanded serologic panel on nucleocapsid (infection-induced antibody) and E484K mutated RBD (delta variant, immune evasive) (E) Nucleocapsid IgG normalized OD at 0 (pink), 6 (green), and 12 months (blue). (F) Normalized OD of Nucleocapsid IgG in spike seropositive unvaccinated and vaccinated participants. Vaccine dose 0 (white) x̃ = 17.04, IQR = 53.50; dose 1 (light blue) x̃ = 13.43, IQR = 28.20; dose 2 (dark blue) x̃ = 11.58, IQR = 14.13. Fully vaccinated with no prior infection (light green) x̃ = 10.80, IQR = 10.62; with known prior infection (dark green) x̃ = 53.79, IQR = 45.32. (G-I) Correlation of antigen OD with other antigens at (G) 0 months, (H) 6 months, and (I) 12 months. Data are distribution or median (x̃) and interquartile range (IQR).

## DISCUSSION

In this study, we observed SARS-CoV-2 seroreactivity in a U.S. population over a 12-month period, including early on in the COVID-19 pandemic, as well as the time directly preceding and succeeding initiation of vaccine rollout. Overall, we saw a robust increase in seropositive participants at 6 months, followed by a further increase at 12 months. While the baseline and 6-month seroprevalence values were likely mostly due to natural infection, it is expected that the vaccine rollout played a significant role in the increase in seropositivity at 12 months. While smaller serosurveys have identified significant trends among small populations, these trends may not be applicable in larger populations (*35, 36*). In this study we utilized a large number of samples from the United States and additionally screened for antibodies against a wide number of antigens to allow for identification of trends in seroprevalence for several different antigens to overcome some of the limitations of smaller serosurveys. This includes assessing levels of anti-S and anti-RBD IgG, IgA, and IgM levels. We also assessed the effect of the E484K mutation within the RBD on antibody recognition to assess the dynamics of mutation-specific seroprevalences. As vaccine rollout occurred, we then analyzed levels of anti-Nuc antibodies in our seropositive samples to delineate between recently infected and non-recently infected seropositive samples. As a result of our serosurvey, we were able to assess broad trends across the entire United States to identify trends and variations based on demographic and socioeconomic factors. Such variations could identify populations who were more vulnerable to infection within these timepoints and inform future pandemic response.

When evaluating the seropositivity changes, we saw several inversions of patterns identified at the 6-month timepoint when compared to 12 months. An obvious variable to consider from these shifts is vaccinations. We saw more individuals with healthcare coverage seropositive for spike antibodies at 12 months, but more nucleocapsid positive individuals in the group that did not have healthcare coverage. Furthermore, there was a larger proportion of individuals with healthcare that received at least 1 vaccination (74.2%) in comparison to those without healthcare (53.0%) at 12 months. The same pattern was true for those that had graduated college (86.2% at least one vaccination) versus those that had graduated high school (75.5%) of those that returned all three samples. In individuals from rural populations, seropositivity was slightly above the overall estimate at 6 months (38.0%) but much lower at 12 months (64.3%). Both antibodies against nucleocapsid and spike were lower in rural populations, suggesting that lower seropositivity could be both as a result of lower vaccination and lower infection rates. The prevalence of severe and complicated SARS-CoV2 infections was noted to be increased in rural areas compared to urban areas (*37*), and slower-developing immunity in these populations may have contributed to this over the course of the pandemic.

Such studies on distinct timeframes (e.g., leading up to commencement of vaccination efforts) during a pandemic can be critically important for evaluation of future viruses with pandemic potential. These data represent a critical portion of the pandemic response that must be analyzed allowing for a window into the timeframe between which there were no vaccines available up until widespread rollout of vaccines. Furthermore, trends in these data help to identify methods of interpreting population-wide susceptibility to future waves of emergent respiratory viruses. The unique nature of having many individuals in a longitudinal serosurvey allows for both population-level weighted estimates, and viewing of different phenomena within the study group, allowing for both public health and individual immunologic pattern evaluation between participants. Not only do these data provide a deeper insight into the dynamics of pathogen spread early in the pandemic, during critical times for public health efforts in mitigating disease spread, it also provides a unique dataset for downline analysis and interpretation.

As with any clinical study there are several limitations to consider that may affect interpretation of data. Antibody waning is a phenomenon that has been described for both vaccination-induced and infection-induced antibodies and has been shown for both S and N antibodies (*38, 39*). Nucleocapsid antibody waning has particularly been emphasized in serological studies, as waning has been reported 5-12 months after infections (*32*). While nucleocapsid waning has been reported to begin as early as 5 months, detectable nucleocapsid antibodies have been reported to persist for over 200 days after infection (*32, 40, 41*). Assays with thresholds determined by a healthy population may not adequately account for variance in background reactivity among patients with different exposure histories or ages. While our overall seroprevalence estimates rely on integrating the findings of two antigens to increase specificity (*6*), the use of only one analyte (Nuc-IgG) to determine infection rates may result in an over-estimate of infections due to slightly decreased specificity (99.01%, 96.49 - 99.88; **Supplemental Figure 1**) and increased chances for error. In terms of the study population, we observed different rates of attrition from certain groups, yielding a skewed population in comparison to the first representative study. Furthermore, the initial sample population was chosen from a convenience sample, so consequent studies from this population have similar biases discussed in our first timepoint study. Because our study focuses on major trends within a large population, we may lose more detailed evaluations from more focused groups within this population.

In conclusion, our data details the dynamics of SARS-CoV-2 seroprevalence within a critical timeframe of the pandemic - the initial spread of SARS-CoV-2 across the US immediately preceding widespread vaccine availability, and immediately following vaccine rollout. As a result, we noted key trends in seroprevalence for the Spike protein, the RBD of the Spike protein, and the nucleocapsid protein. Such trends could identify populations that are vulnerable to infection in future respiratory viral pandemics, as well as critically evaluate the protective effect of vaccination within these populations. These follow-up studies from our initial study on infection-induced antibody prevalence and reactivity to immune evasive variants provides useful insights into potential spread of emerging respiratory pathogens.

## MATERIALS AND METHODS

### Study design

This clinical study was performed as a continuation of the initial national serosurvey study PMID: 34158410 (ClinicalTrials.gov NCT04334954) (*11, 42*). This study was approved by the NIH Institutional Review Board and conducted in accordance with the provisions of the Declaration of Helsinki and Good Clinical Practice guidelines. All participants provided verbal informed consent before enrollment.

### Blood sample collection

Blood was collected via remote dried blood sampling as per previously published (*11*). Briefly, participants were mailed a Mitra microsampling kit (Neoteryx, Torrance, CA) with four 20 μl sampling tips that collected blood via fingerstick sampling. Samples were shipped overnight directly to the laboratory at the National Institutes of Health, logged, and stored at - 80°C until elution. Samplers that were not filled completely (< 20 μl) as determined by visual inspection, or whose samples were contaminated due to disruption in packaging during shipping were discarded and not included in analysis.

### Protein production

Spike trimers and spike receptor binding domain (RBD) constructs were synthesized and purified as previously published (*43*, *44*). Constructs for full length SARS-CoV-2 Wuhan Nucleocapsid (N, amino acids 1-421) and a C-terminal fragment (N-CTD, amino acids 247-364) were generated as E. coli optimized Gateway Entry clones by ATUM, Inc. The C-terminal (CTD) fragment was preceded by a tobacco etch virus (TEV) protease cleavage site, ENLYFQG. Entry clones were transferred to pDest-527 (Addgene #11518) using Gateway LR recombination per the manufacturer’s protocols (Thermo Fisher Scientific). Final clones contained an aminoterminal His6 tag for purification. Proteins were expressed in 2 liters of E. coli using dynamite broth as described in Taylor, et al. Cell pellets were lysed in 20 mM HEPES pH 7.3, 300 mM NaCl, 1 mM TCEP using Microfluidizer M-110EH by 2 passes at 10,000 psi. Lysates were clarified by centrifugation at 8000 x g for 90 minutes in a JLA 10.500 rotor and frozen at - 70C. Thawed lysate was filtered using a Sartorius Sartopore 2 Capsule, 0.2 µm, 300 cm2, and adjusted to 35 mM imidazole and loaded onto a IMAC column. The equilibration buffer (EB) for the column was 20 mM HEPES, pH 7.3, 3000 mM NaCl, 1 mM TCEP, 35 mM imidazole. The column was washed to baseline with 2 column volumes (CV) of buffer EB and proteins were eluted with a 10 CV gradient of buffer EB from 35 mM to 500 mM imidazole. Elution fractions were analyzed by SDS-PAGE and Coomassie-staining. For N-CTD purification, positive fractions were pooled, dialyzed against 4 liters of dialysis buffer (DB) of 20 mM HEPES, pH 7.3, 300 mM NaCl, 1 mM TCEP in the presence of His6-TEV protease at 6% (v/v), for 2 hours at room temperature and then dialyzed overnight at 4°C. Sample was loaded on an IMAC column equilibrated with DB. After washing to a baseline absorbance, protein was eluted with a 5 CV gradient from 0-50 mM imidazole. Pooled protein was concentrated and applied to a Superdex S-75 26/60 column (Cytiva) equilibrated in 20 mM HEPES, pH 7.3, 150 mM NaCl, 1 mM TCEP. Chromatography was analyzed by SDS-PAGE gel and positive fractions were pooled and concentrated for final protein.

For full length N protein, the Tev digestion step was omitted and replaced by ion exchange chromatography using a HiPrep Q XL 16/10 column. Protein was dialyzed from the IMAC step into 20 mM HEPES, pH 7.3, 100 mM NaCl, 1 mM TCEP and loaded onto a column equilibrated in the same buffer. Protein was eluted using a gradient of buffer from 100 mM to 1.0 M NaCl, and positive fractions were pooled, concentrated, and subjected to size exclusion chromatography on a Superdex S-75 column as noted above for N-CTD. Final proteins were validated by SDS-PAGE, electrospray mass spectrometry, and analytical size exclusion chromatography. Nucleocapsid CTD was selected for serology due to protein aggregation seen with the full-length construct.

### Enzyme-linked Immunosorbent Assay

To assess for seropositivity, a previously established protocol was optimized to a 384 well plate format (*6*). Briefly, one (1) microsampler was loaded into the well of a 1 mL deep 96 well plate (ThermoFisher) that was then stored at 4 degrees until processing and the remaining three were placed in an 0.5 mL Eppendorf tube and stored at -80°C as previously mentioned. Twenty-four (24) to 48 hours prior to analyzing the sample, 400 μl of elution buffer (400 μl 1x PBS (GIBCO) + 1.0% BSA (Sigma) + 0.5% Tween20 (Sigma)) was added to the sample to create an eluate.

All steps of the ELISA apart from addition of coating solution and sample were done using a BioTek EL406. To prepare plates, 50 μl of coating solution consisting of either 1 μg/mL of Spike (trimer), 2 μg/mL of RBD/RBD Variants (ex-E484K), or 2 μg/mL of Nucleocapsid in 1x PBS were added to 384 well Nunc Maxisorp plates (Fisher, 464718) and incubated for 16 hours at 4 degrees Celsius. Wells were washed 3 times with Wash buffer consisting of 0.05% Tween20 (Sigma) in 1x PBS before adding 100 μl of blocking buffer, consisting of 5% Non-Fat Dried Milk and 0.05% Tween20 in 1x PBS. Plates were incubated with blocking buffer for 2 hours.

During incubation, sample eluate was added either 1:10 or 1:1,000 in sample buffer consisting of 5% Nonfat Dried Milk in 1x PBS. As a positive control, a standard curve consisting of a mix of IgA, IgM, and IgG anti-SPIKE recombinant antibodies (GenScript Custom IgA, Thermo Cat #A02046, and Thermo Cat #A02038) was diluted at a concentration of 1:100, 1:500, 1:1000, 1:2500, 1:5000, and 1:10,000 in pre-pandemic pooled serum to account for any background signal due to serum. Recombinant antibody standard curve samples were then added 1:400 μl to blocking buffer, as previously described. For nucleocapsid plates, a standard curve of the same dilutions was performed using an anti-Nucleocapsid Antibody (Fisher, #MA535942).

After 2 hours of blocking, plates were washed three times prior to addition of sample. Fifty (50) μl of sample was added per well. Incubation with sample was one hour, during which secondary antibody buffers were prepared. Secondary antibody solution consisted of a 0.25 μg/mL solution of IgA, IgM, or IgG Secondary Antibody (Fisher, A18787, A18841, A18811) in blocking buffer. Fifty (50) μl of secondary antibody solution was added to corresponding plates and incubated for an hour. Plates were washed with wash buffer three times prior to addition of 30 μl of substrate TMB (Fisher, 34029) and 30 μl of stop solution (Fisher, SS04). Absorbance at 450 and 650 nm was recorded using the BioTek Epoch2 plate reader. Assay stability and range are shown in **Supplemental Figures 3-4**.

### Statistical Analysis

The study consisted of a baseline survey with 8,058 participants who provided complete data in Kalish et al. (11), followed by a six- and twelve-month follow-up involving a web survey and home blood microsampling kit. Respondents were defined as participants who returned their blood samples, resulting in 4,562 and 4,226 respondents at months 6 and 12, respectively. The statistical analysis proceeded in three steps. Firstly, kernel-weighted propensity-score matching pseudoweights (KW) were constructed, as described in Wang et al. (2022) (*45*), based on sixteen demographic and health-related questions (see Table 1 in Kalish et al., 2021 (*11*)). The distribution of these questions in the quota sample was matched to those of the BRFSS survey, a large probability-based national survey by KW-weighting (*46*). Secondly, KW were adjusted using weighting based on the propensity of responding to the follow-up surveys, in order to account for potential nonresponse bias. This was achieved by estimating the response propensity using important covariates collected in the baseline survey that may be related to both response propensity and seropositivity. Finally, nonresponse-adjusted KW-weighted estimators were further adjusted to account for sensitivity and specificity (43). Confidence intervals were then calculated for the final seroprevalence estimates, accounting for variabilities due to KW-weighting, nonresponse propensity weighting, and the sensitivity and specificity adjustment (*47*).

Additional descriptive analyses plotted raw serology data at 6 and 12 months, and antibody dynamics longitudinally at baseline, 6, and 12 months, along with 95% confidence intervals (*48*). Participants were included for this analysis if they had all three measurements available.

## Data Availability

All data produced in the present work are contained in the manuscript.

## ACKNOWLEDGEMENTS

The authors would like to acknowledge and thank all participants in this study without whom the research would not be possible. This research was supported, in part, by the Intramural Research Program of the NIH, including the National Institute for Biomedical Imaging and Bioengineering, the National Institute of Allergy and Infectious Disease, and the National Center for Advancing Translational Sciences, and the National Cancer Institute. This project has been funded, in part, with Federal funds from the National Cancer Institute, NIH, under contract number HHSN261200800001E, 75N91019D00024, Clinical and Translational Science Awards Program grants UL1TR003096 (UAB) and UL1TR001857 (University of Pittsburgh). Thanks to the University of Pittsburgh Clinical and Translational Science Institute (CTSI): J. Avolio, M. Archila, L. Bash, K. Carroll, S. Clayton, M. Cristinziano, J. Crnkovic, K. Daw, C. Derrow, T. Ditter, C. Fascetti, M. Glenn, E. Gyurisin, J. Huwe, S. Igwe, N. Jones, B. Kalchthaler, M. Kienholz, D. Limpert, T. Malay, D. Mathias, S. Mathias, E. Miller, J. Mullins, A. Mykita, A. Packard, B. Petersen, M. Phillips, C. Rush, E. Shepherd, S. Shetty, N. Siripong, A. Socci, L. Stearns, H. Strausser, A. Thompson, S. Ugbomah, K. Underwood, L. Yasko) and University of Pittsburgh Information Technology (D. McGaughey, S. Ritzman, T. Smith) for their contributions to this work. <u>Disclaimer:</u> The content of this publication does not necessarily reflect the views or policies of the Department of Health and Human Services, nor does mention of trade names, commercial products, or organizations imply endorsement by the U.S. government. The NIH, its officers, and employees do not recommend or endorse any company, product, or service.

