## Supplementary Materials for "Dynamics of SARS-CoV-2 Seroprevalence in a Large US population Over a Period of 12 Months"

Karkanitsa, Li, Valenti et al.

**This PDF file includes:**

Figs. S1 to S5

**SUPPLEMENTARY FIGURES AND LEGENDS**

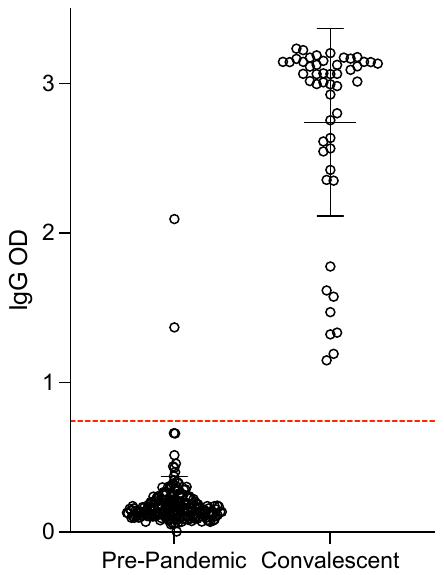

**Supplemental Figure 1: Nucleocapsid assay controls.** Pre-pandemic and convalescent controls for nucleocapsid IgG assay. Red line = positivity cutoff (mean + 3SD).

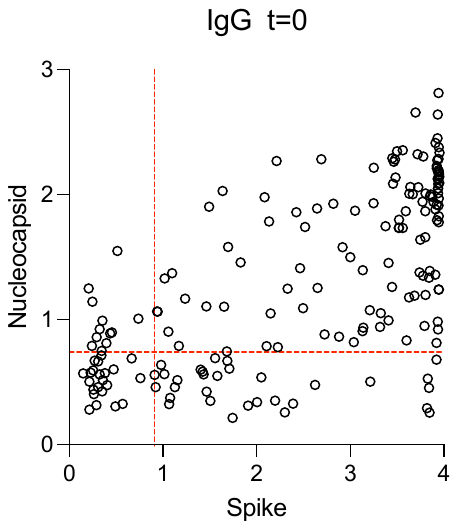

**Supplemental Figure 2: Nucleocapsid versus Spike IgG samples at baseline**. Nuclecapsid IgG levels in seropositive individuals from the baseline measurement. Red = positivity cutoff. Data = raw optical density (OD).

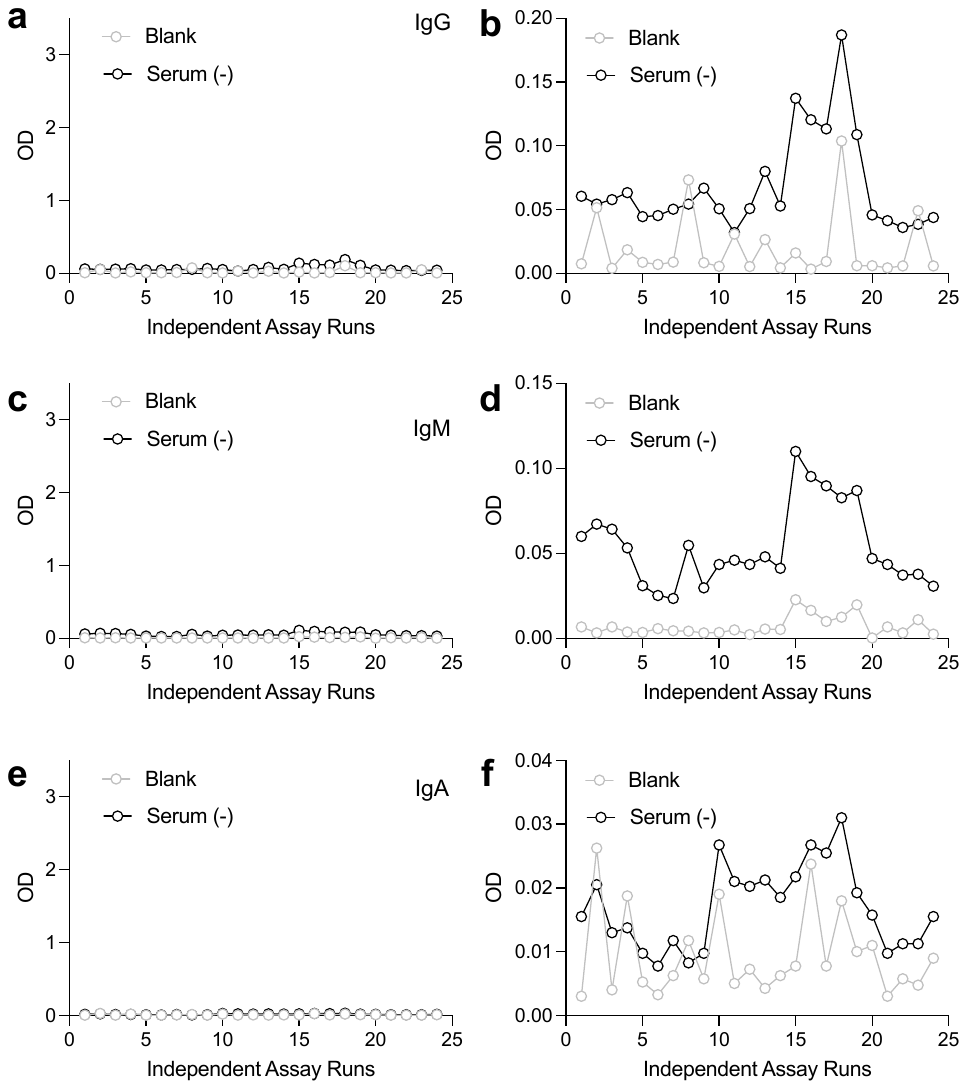

**Supplemental Figure 3: Anti-Spike ELISA assay stability of negative controls**. Grey = blocking buffer alone (blank); Black = pre-pandemic seronegative serum control. (a-b) IgG (c-d) IgM, (e-f) IgA. Graphs in b,d, and f are truncated Y-axes to show variation.

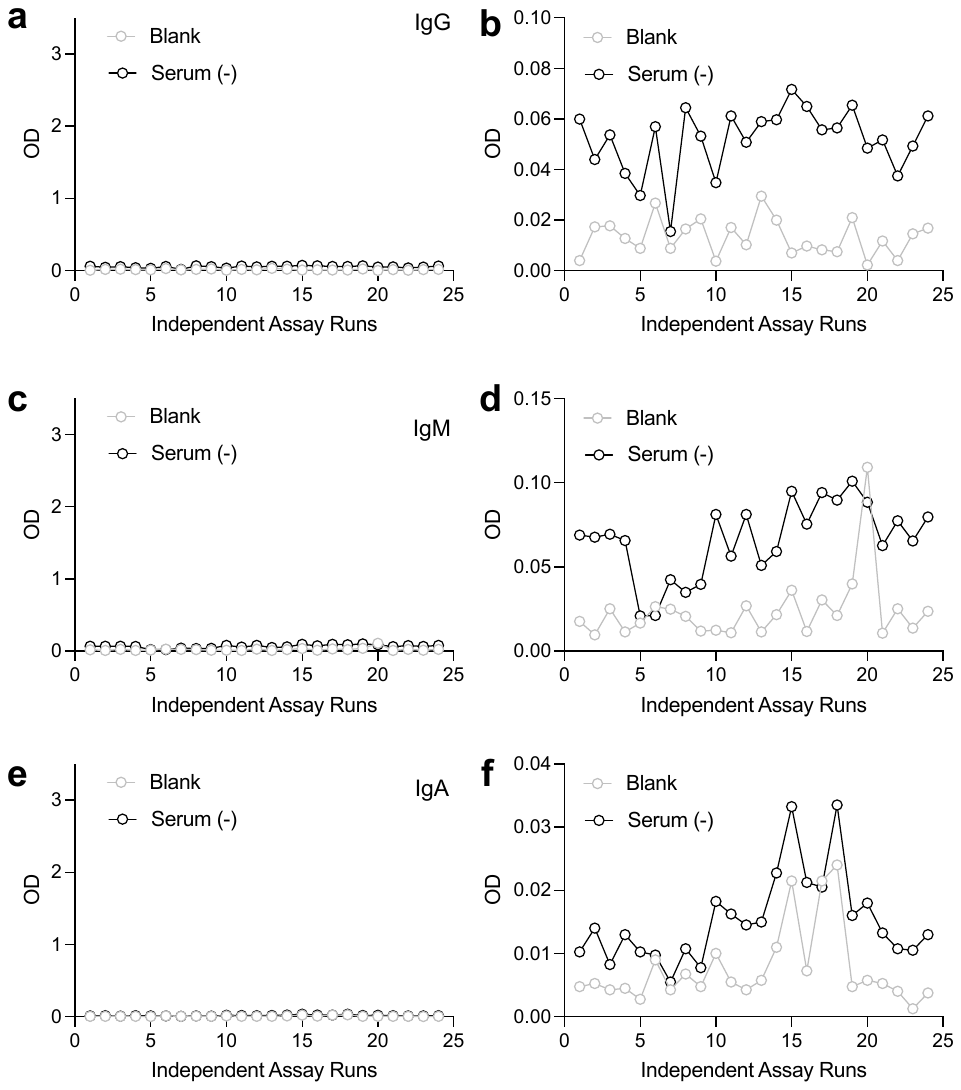

**Supplemental Figure 4: Anti-RBD ELISA assay stability of negative controls**. Grey = blocking buffer alone (blank); Black = pre-pandemic seronegative serum control. (a-b) IgG (c-d) IgM, (e-f) IgA. Graphs in b,d, and f are truncated Y-axes to show variation.

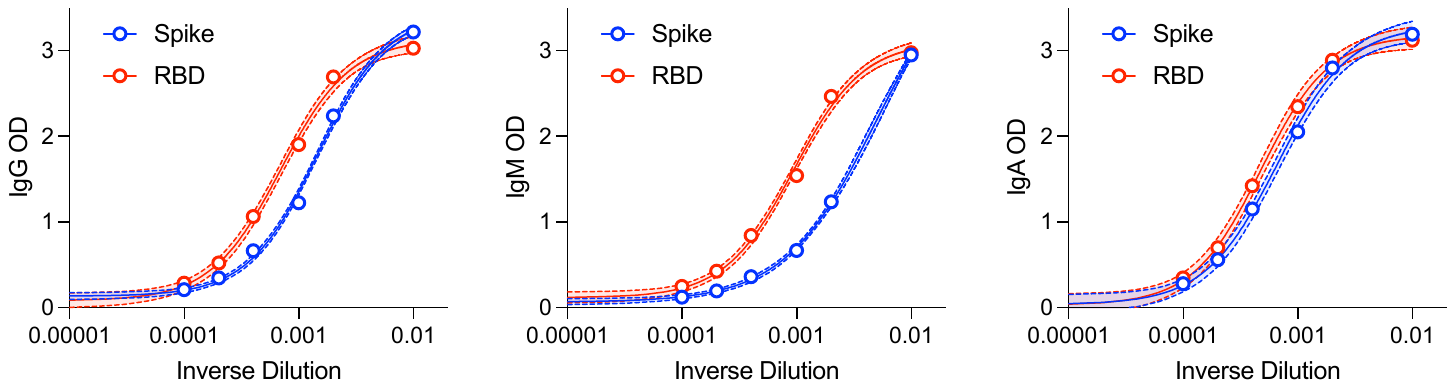

**Supplemental Figure 5: Positive control antibody titration curves.** Recombinant monoclonal antibodies spiked into seronegative serum for assay range display. Red = RBD, Blue = Spike. Data are Sigmoidal 4 parameter logistic regression with 95% confidence intervals.
